# Antimicrobial resistance profiling and phylogenetic analysis of *Neisseria gonorrhoeae* clinical isolates from Kenya in a resource-limited setting

**DOI:** 10.1101/2020.09.18.20185728

**Authors:** Meshack Juma, Arun Sankaradoss, Redcliff Ndombi, Patrick Mwaura, Tina Damodar, Junaid Nazir, Awadhesh Pandit, Rupsy Khurana, Moses Masika, Ruth Chirchir, John Gachie, Sudhir Krishna, Ramanathan Sowdhamini, Omu Anzala, Meenakshi S Iyer

## Abstract

**Background:** Africa has one of the highest incidences of gonorrhoea, but not much information is available on the relatedness with strains from other geographical locations. Antimicrobial resistance (AMR) in *Neisseria gonorrhoeae* is a major public health threat, with the bacteria gaining resistance to most of the available antibiotics, compromising treatment across the world. Whole-genome sequencing is an efficient way of predicting AMR determinants and their spread in the human population. Previous studies on Kenyan gonococcal samples have focused on plasmid-mediated drug resistance and fluoroquinolone resistance using Illumina sequencing.

Recent advances in next-generation sequencing technologies like Oxford Nanopore Technology (ONT) have helped in the generation of longer reads of DNA in a shorter duration with lower cost. However, long-reads are error-prone. The increasing accuracy ofbase-calling algorithms, high throughput, error-correction strategies, and ease of using the mobile sequencer in remote areas is leading to the adoption of the MinION sequencer (ONT), for routine microbial genome sequencing.

**Methods:** To investigate whether MinION-only sequencing is sufficient for diagnosis, genome sequencing and downstream analysis like inferring phylogenetic relationships and detection of AMR in resource-limited settings, we sequenced the genomes of fourteen clinical isolates suspected to be N. gonorrhoeae from Nairobi, Kenya. The isolates were tested using standard bacteriological methods for identification, interpreted using analytical profile index and antibiotic susceptibility tests had indicated ciprofloxacin and gentamycin resistance. Using whole-genome sequencing, the isolates were confirmed to be cases of N. gonorrhoeae (n=12), Additionally, we identified reads from N. meningitidis (n=2) and both of N. gonorrhoeae and Moraxella osloensis (n=3) in the sample (co-infections) respectively, which have been implicated in sexually transmitted infections in the recent years. The near-complete N. gonorrhoeae genomes (n=10) were analysed further for mutations/factors causing AMR using an in-house database of mutations curated from the literature. We attempted to understand the basis of drug resistance using homology modelling of AMR proteins, using known structures from other bacteria.

**Results:** We observe that Ciprofloxacin resistance is associated with multiple mutations in both *gyrA* and *parC*. We identified mutations conferring tetracycline (*rpsJ*) and Sulfonamide (*folA*) resistance in all the isolates and plasmids encoding beta-lactamase and *tet*(M) were identified in almost all of the strains. Phylogenetic analysis clustered the nine isolates into clades containing previously sequenced genomes from Kenya and countries across the world.

**Conclusion:** Here, we demonstrate the utility of mobile DNA sequencing technology supplemented with reference-based assembly in sequence typing and elucidating the basis of AMR. Bioinformatics profiling to predict AMR can be used along with routine AMR susceptibility tests in clinics. The workflow followed in the study, including AMR mutation dataset creation and the genome identification, assembly and analysis, can be used for the genome assembly and analysis of any clinical isolate. Further studies are required to determine the utility of real-time sequencing in the outbreak investigations, diagnosis and management of infections, especially in resource-limited settings.

**Data availability:** The raw reads generated for this study have been deposited in BioProject Accession: PRJNA660404 (https://www.ncbi.nlm.nih.gov). The biosample details are available under the ids SAMN15960547, SAMN15960548, SAMN15960549, SAMN15960550, SAMN15960551, SAMN15960552, SAMN15960553, SAMN15960554, SAMN15960555. The genomes and annotation files are available under the bioproject.

## Introduction

Gonorrhoea is one of the most common sexually transmitted infections (STIs), which is caused by *N. gonorrhoeae* (gonococci). It is estimated to have affected 87 million people (between the ages 15 to 49) worldwide, 11.4 million in the African Region, in 2016^1^. Gonorrhoea can cause epididymitis in men and pelvic inflammatory disease in women, which can result in infertility and ectopic pregnancies. It is also implicated in neonatal ophthalmia, which can result in blindness and increased acquisition and transmission of other STIs^2^. WHO included *N. gonorrhoeae* in its list of AMR “priority pathogens” in 2017^3^ and has projected for a 90% reduction in global gonorrhoea incidence by 2030^1^. Although *N. gonorrhoeae* infections are treated with antibiotics, gonococci are now resistant to most antibiotics and despite two clinical trials, no vaccines are available^4^. Increasing antimicrobial resistance threatens effective treatment and control.

Many clinical samples are developing resistance to extended-spectrum cephalosporins (ESCs) and azithromycin, which are used as the last options of the first-line therapies of gonorrhoea. In many countries, ceftriaxone and azithromycin dual therapy is being used^5^. However, with many isolates showing increased MICs to both these antibiotics, this is not viable as a long-term treatment^6^. Many countries in the African region have reported decreased susceptibility to drugs like ceftriaxone, azithromycin, ciprofloxacin^4^. Whole-genome sequencing (WGS) allows for timely detection and elucidation of AMR determinants in bacteria^7^. Third-generation sequencing has many advantages, like long read lengths, short time, and reduction of bias introduces through amplification by PCR-steps^8^. WGS has been used to investigate quinolone-resistant gonorrhoea outbreaks^9,10^ in Kenya. It has also been used to analyse isolates resistant to ciprofloxacin, azithromycin and cefixime in other countries^11–13^.

MinION, a mobile sequencing device from ONT (Oxford, UK), sequences DNA by monitoring the transfer of individual DNA molecules through various types of pores, resulting in very long and unbiased sequence reads, as there is no amplification or chemical reactions used during sequencing^11^. The mobile nature of the MinION offers many advantages and the sequencer has been evaluated for use in resource-limited settings and on the International Space Station (ISS)^14,15^. MinION has also been shown to be effective for diagnostics, high-quality assemblies, and AMR surveillance in bacteria. However, the high error rate for the sequencer limits its use. Golparian and co-workers sequenced fourteen clinical isolates of *N. gonorrhoeae* and evaluated multiple methods for genome assemblies using MinION with Illumina reads^11^. Street and co-workers used MinION to obtain a *de novo* assembly of *N. gonorrhoeae* isolated from patient urine samples^16^. Zhang and co-workers showed that AMR-profiling results for *N. gonorrhoeae* were comparable when using only MinION-based assemblies or MinION-Illumina hybrid assemblies.

In a previous study, 22 multi-drug resistant gonococcal isolates from heterosexual patients (2 females and 20 males) were sequenced using Illumina MiSeq for understanding *gyrA* and *parC* mutations in ciprofloxacin resistance^10^. Cehovin et al sequenced 103 genomes from 73 patients from coastal Kenya, mainly men who had sex with men, using Illumina HiSeq and analyzed the plasmids conferring antibiotic resistance^17^. These studies in Kenya have used the Illumina platform and mainly focused on isolates from male patients. We sequenced nine isolates derived from women who visited STI clinics in Nairobi between 2012-2017, using only MinION sequencing in Kenya and evaluated methods for assembly and AMR profiling of genome. We have also carried out phylogenies with previously deposited sequences in PubMLST, from Kenya, and other countries. Here we assess the possibility of using MinION data alone for genome assembly and comparative analysis.

## Materials and methods

### Bacterial isolates and culture conditions

*N. gonorrhoeae* isolates were derived from high vaginal swabs collected between January 2012 to December 2017. The stocked organisms were stored at -70ºC and retrieved and cultured on Modified Thayer-Martin agar. The plates were incubated at 37°C and 5% carbon dioxide (under raised CO2) as per the standard protocol.

Isolates were identified by Gram stain, Catalase test, Oxidase test, and carbohydrate-utilization studies of glucose, maltose, fructose and sucrose, and confirmed by analytical profile index testing kit (API NH, bioMérieux). All gonococcus isolates were re-stocked in 20% glycerol-tryptic soy broth for long-term storage at −70°C for external quality control, sequencing and molecular characterization. Standard *N. gonorrhoea* isolates i.e WHO K, WHO P, WHO O, WHO R, WHO M^18^ were used to test the media for viability and colonial characteristics. Phenotypic characterization was performed by characterizing lactamase production determined using nitrocefin solution. In the present study, clinical isolates from nine patients were sequenced (Supplementary Table 1).

### Antibiotic susceptibility testing

Ciprofloxacin, Spectinomycin, Penicillin, Tetracycline, Cefixime, Ceftriaxone, Erythromycin and Azithromycin strips were used to set E-test according to the Clinical and Laboratory Standards Institute (CLSI). Antimicrobial gradient diffusion (E-test) method was used to determine the minimum inhibitory concentrations (MIC) of *N. gonorrhoeae* using WHO reference ranges, interpretation of susceptibilities for selected antimicrobial agents as recommended by CLSI (Supplementary Table 2).

### Isolation of genomic DNA

All the colonies from Thayer-Martin agar (37°C in a humidified 5% CO2 environment for 36 hours) with the phenotypic characteristics of *N. gonorrhoeae* were picked up to create a culture suspension in 1X PBS. The genomic DNA was isolated using the Qiagen DNA isolation kit using manufactures specifications. Extracted DNA was purified using AMPure XP beads (Beckman Coulter, UK), eluted in 50 μl of nuclease-free water, and quantified using a QiaXpert (Qiagen). Pipetting was minimized to reduce shearing of the DNA prior to sequencing.

### ONT library preparation and MinION sequencing

DNA library preparation for Nanopore sequencing was carried out using Ligation Kit SQKLSK109 (ONT). Fragmented DNA was repaired and dA-tailed using the NEBNext FFPE DNA Repair Mix and NEBNext Ultra II End Repair/dA-Tailing Module (New England BioLabs). An individual barcode was added to dA-tailed DNA by using the barcoding extension kit EXPNPB104 in accordance with the ONT protocols with NEB Blunt/TA Ligase Master Mix (New England BioLabs). Each barcoded DNA was pooled in equimolar amounts, and an adaptor was attached using the NEBNext Quick Ligation Module (New England BioLabs). The MinION flow cell and reagents were shipped from Bangalore, India, to Nairobi at +4°C. The number of active pores was checked before loading. The samples were pooled using equimolar pooling, the library was loaded into the SpotON flowcell R9.5 (FLO-MIN106), and sequencing was carried out on MinKNOW using the 48-hour script.

### Basecalling, Read trimming and processing

Guppy (v 3.2.2) (ONT) was used for basecalling of fast5 files. Reads with a mean qscore (quality) greater than 7 and a read length greater than 500bp were used and trimmed for adaptor sequences and barcodes using qcat (v1.1.0) (https://github.com/nanoporetech/qcat) from ONT. The files corresponding to each barcode (sample) were analyzed as follows.

### Assembly and assessment

The selected raw reads were corrected, trimmed and assembled using Canu (v1.8) (https://github.com/marbl/canu) using the parameters genomeSize=2.1m and minReadLength=500^19^. Minimap (v 2.17) (https://github.com/isovic/racon) and Miniasm (v0.3) (https://github.com/lh3/miniasm) were used for the self-mapping of the raw reads and the concatenation of the alignments to get the *de novo* assembly, respectively^20^. Two rounds of error-correction of this draft assembly were carried out with Racon (v1.4.3) (https://github.com/isovic/racon) using raw reads^21^. The above assemblies were polished using the raw nanopore reads with Nanopolish (v0.11.1) (https://github.com/jts/nanopolish)^22^. We also carried out a guided genome assembly using reference genome *N. gonorrhoeae* FA 1090 (NCBI:txid485). The trimmed raw reads were aligned to the reference genome using the bwamem option from bwa (v0.7.12) (http://bio-bwa.sourceforge.net/)^23^. Samtools (v1.9) (http://www.htslib.org/) was used for deriving the bam alignment file and the consensus assembly (bcftools), after normalizing the indels and variant calling^24^. The options bcftools filter --IndelGap 5 and bcftools consensus -H ‘A’ were used for deriving the consensus assembly. Mummer (v3.9.4) (http://mummer.sourceforge.net/)^25^ and bedtools (v2.25.0) (https://bedtools.readthedocs.io/en/latest/)^26^ were used for calculating the correspondence of the assemblies with the reference genome, genome coverage and sequencing depth. The base positions with zero coverage were extracted using awk commands and bedtools maskfasta option was used to derive the draft genomes, missing nucleotides were marked in the assembly as N.

### Genome annotation

The gene prediction and annotation were carried out using the RAST server (https://rast.nmpdr.org/) using the reference genome for guiding the gene prediction^27^.

### Genome-based MLST analysis

Gene profiles for sequence typing of *N. gonorrhoeae* were downloaded from different databases like NGSTAR (*N. gonorrhoeae* Sequence Typing for Antimicrobial Resistance) (https://ngstar.canada.ca/)^28^, PubMLST (Public databases for multi-locus sequence-typing) (https://pubmlst.org/)^29,30^ and NGMAST (*N. gonorrhoeae* multi-antigen sequence typing) (http://www.ng-mast.net/)^31^. NGMAST uses two highly polymorphic loci, the outer-membrane porin (*por*) and transferrin binding protein β-subunit *tbpB*, for sequence typing. NGSTAR uses variants in seven antimicrobial resistance determinants (*penA*, *mtrR*, *porB*, *ponA*, *gyrA*, *parC* and 23S rRNA) to three classes of antibiotics (cephalosporins, macrolides and fluoroquinolones) and PubMLST uses seven housekeeping genes (*abcZ*, *adk*, *aroE*, *fumC*, *gdh*, *pdhC*, and *pgm*) for sequence typing. Profiles were created and blastn (E-value 10-7) from standalone BLAST+ (https://ftp.ncbi.nlm.nih.gov/blast/executables/blast+/)^32^ was used to assign the genes to the different alleles for MLST genes.

### Identification of mutations causing AMR

Around 85 mutations in 14 genes reported to be involved in AMR were checked for in the sequenced strains, including promoter mutations resulting in overexpression of efflux proteins^33^. Additionally, we checked for the presence of four drug efflux pumps, two genes from conjugative plasmids and plasmid-mediated AMR determinants (Supplementary File). Mutations were manually screened for, in the protein sequences identified using the RAST server in all the genomes. Sequencing depth at the corresponding gene loci was calculated using a custom shell script. We compared our results with that of two publicly available databases for AMR, CARD database and PathogenWatch^34,35^. Resistance genes from plasmids were identified using blastn (E-value 10^-7^) against NCBI Bacterial Antimicrobial Resistance Reference Gene Database (https://www.ncbi.nlm.nih.gov/bioproject/PRJNA313047/)^36^.

### Homology modelling

To understand the basis of antibiotic resistance, wild-type and mutant proteins implicated in AMR, were modelled using templates from other bacteria. Co-ordinates of heteroatoms like antibiotics and DNA are present in the templates used. Hence, the heteroatom modelling module from Modeller (v9.23)^37^ was used to build a multi-chain model with symmetry restraints. Clustal Omega (https://www.ebi.ac.uk/Tools/msa/clustalo/) was used for the alignment of query protein sequences with the template protein sequence, with manual correction of the alignment^38^. The residues in different chains were separated using ‘/’ and the symbol ‘.’ was used to indicate the ligand molecules, in the alignment.

### Multiple-sequence alignment and phylogenetics

Phylogenetic tree analysis was carried out using 123 previously sequenced *N. gonorrhoeae* strains and 130 strains from different countries across the world, standard WHO strains and the reference genome. The genome analysis module from Bacterial Isolate Genome Sequence Database (BIGSdb) (https://pubmlst.org/software/database/bigsdb/) was used to generate neighbour net tree structure based on distance matrices^30^ using core genome alignments. Phylogenetic trees were derived using the Neighbour Joining (NJ) module using SplitsTree. The trees were annotated with details like WHO geographical region and antibiotic sensitivity using iTOL.

The workflow for reference-based assembly used in the study has been illustrated in Figure 1a, and the subsequent analysis workflow has been shown in Figure 1b. The workflow for the *de novo* assemblies has been shown in Supplementary Figure 1.

**Figure 1:**
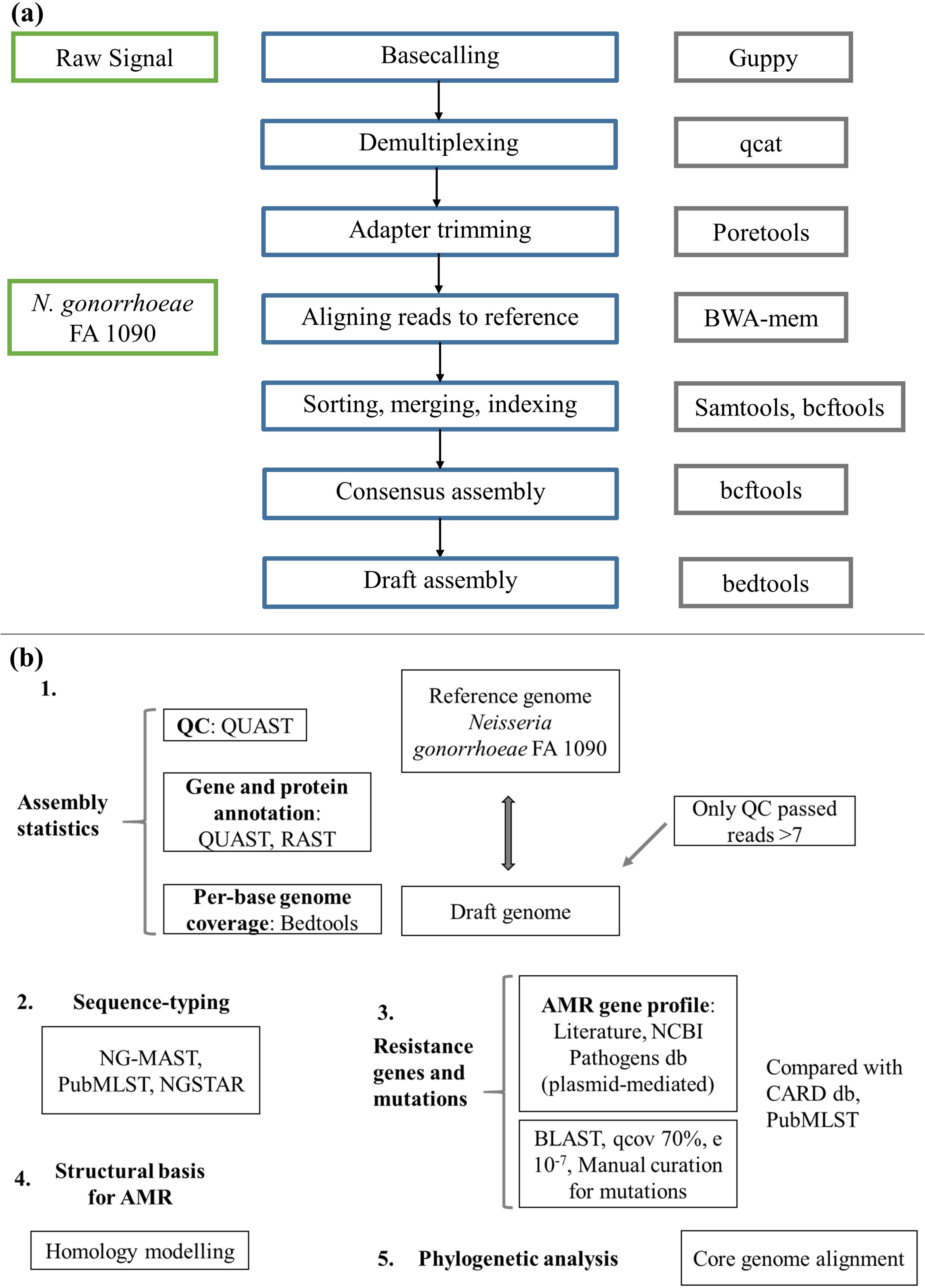
The work-flow for the reference-based assembly (a) and downstream analysis (b) of the nine genomes sequenced in the study. We carried out a reference-based genome assembly and used RAST for genome annotation. For sequence-typing, we carried out BLAST searches with alleles derived from PubMLST and a set of AMR determinants was identified through literature and was used for screening the clinical isolates. Structure modelling of the proteins with mutations was carried out using templates with crystallised antibiotic structures to understand the basis of drug resistance.

## Results

### Overview of sequenced data

We obtained between 534-2387 Mb of reads from the MinION sequencing runs for each sample. The longest reads ranged from 156-406 kb with an average length of 1.5-2.7 kb (Table 1).

**Table 1:** Read and assembly statistics. The number of mapped reads for each genome was obtained using samtools flagstat and the genome assembly statistics were obtained using QUAST. The genome coverage was not affected by the number of reads or the sequencing depth.

| Strain IDs | Genome coverage (%) | Sequencing depth | #mapped reads |
| --- | --- | --- | --- |
| 3 | 99.03 | 523.604 | 308387 |
| 12 | 99 | 377.339 | 216691 |
| 18 | 98.78 | 272.294 | 175050 |
| 57 | 98.81 | 1255.388 | 2272143 |
| 61 | 98.97 | 63.726 | 43553 |
| 100A | 98.68 | 848.073 | 1460162 |
| 240 | 99.59 | 86.3 | 70185 |
| 274 | 98.88 | 240.667 | 180375 |
| 285 | 98.31 | 129.085 | 116342 |
| 298 | 98.67 | 219.616 | 212198 |

### Genome assembly statistics and species identification

Overview of the mappability of the reads from different strains to the reference genome has been depicted in Supplementary Figure 2. Assemblies were compared and selected based on the following criteria: the lowest number of mismatches, misassemblies, contigs, and the highest fraction of genome coverage using QUAST^39^ (Supplementary Table 3a-c). The length of the assemblies and the number of CDS identified were fairly the same across different methods. No plasmid or transposon gene insertions were observed in the genomes assembled *de novo* and the reference-based assembly. The species of the *de novo* assembled genome was confirmed using the Ribosomal Multilocus Sequence Typing rMLST^40^ approach from BIGSdb, PubMLST to identify the causative organism and co-infections. The database uses 53 genes encoding the bacterial ribosome protein subunits (rps genes) for species assignment. We also used blastn with 16S rRNA and 23S rRNA as the queries for species confirmation. The reference-based assembly obtained from bwa-mem and the plasmid sequences assembled using Canu and Minimap were used for further analysis. The plasmid sequences were used for detecting the AMR determinants and no additional downstream analysis was performed on them.

### Sequence-typing of the strains

The alleles used in sequence-typing (cgMLST, NGMAST and NGSTAR) were assigned to the genes from different strains, and we identified few novel alleles (Supplementary Tables 4a-c). The sequencing depth at each of these loci was found to be very high (>50-600) (Supplementary Table 4d). The novel allele sequences have been deposited in the corresponding databases.

### Antibiotic resistance and mutations in AMR genes

The MIC results with six different antibiotics have been provided in Supplementary Table 5.

A dataset of around 89 AMR targets was identified using literature (Supplementary Table 6a). Out of these, 23mutations (in nine genes) were observed in the strains (Table 2). The sequencing depth of *gyrA*, *parC*, *mtrR*, *ponA1*, *penA*, *porB*, *folP*, *rplD* and *rpsJ* genes was high (Supplementary Table 4d and Supplementary Table 6b). Other databases used for AMR profiling identified only a fraction of the mutations we identified (Supplementary Table 6c).

**Table 2:** AMR gene profile of the nine strains. Mutations observed in the studied strains have been listed, mutated residues are indicated in red, mutations identified in this study have been indicated in bold. The resistance profile for the strains as inferred through MIC has been indicated. The references and antibiotic-mutation association has been provided in Supplementary Table 5a.

| Gene | Mutation | Genome |  |  |  |  |  |  |  |  |  |  |
| --- | --- | --- | --- | --- | --- | --- | --- | --- | --- | --- | --- | --- |
|  |  | Reference | 3 | 12 | 18 | 57 | 61 | 100A | 240 | 274 | 285 | 298 |
| *gyrA* | S91F/Y | S | S | F | F | F | S | F | F | S | F | S |
|  | D95N/G/A | D | D | A | A | A | D | A | D | D | A | D |
| *parC* | V68A | V | V | V | V | V | V | V | V | V | A | V |
|  | E91K/G | E | E | G | E | E | E | E | G | E | G | G |
|  | Y104 silent | TAC | TAT | TAT | TAT | TAT | TAT | TAT | TAT | TAT | TAT | TAC |
|  | L131 silent | CTG | CTA | CTG | CTG | CTG | CTG | CTG | CTC | CTG | CTG | CTG |
| *ponA1*<br>(*pbp1*) | L421P | L | P | L | P | P | L | P | P | L | P | L |
| *rpsJ* | V57M | V | M | M | M | M | M | M | M | M | M | M |
| *penA/*<br>*pbp2* | D345<br>insertion | - | D | D | D | D | D | D | D | D | D | D |
|  | F504L | F | L | L | L | L | L | L | L | L | L | L |
|  | A510V | A | V | V | V | V | V | V | V | V | V | V |
|  | A516G | A | G | G | G | G | G | G | G | G | G | G |
|  | P551S/L | P | P | P | S | S | P | S | L | S | S | S |
| *porB1b*<br>(*penb*) | G101K/L/N | K | K | K | K | K | K | K | K | K | K | K |
|  | A102D/N/ | G | G | G | G | G | G | G | G | G | G | G |
|  | G120D/K | G | G | G | D | D | G | D | D | G | G | G |
|  | A121D | A | A | A | D | G | A | G | A | S | S | G |
|  | N122 | N | N | N | N | D | N | S | N | K | K | N |
| *folP* | T66M | T | M | T | T | T | T | T | M | T | M | T |
|  | P68S/L/A/Q | P | P | P | Q | P | Q | P | P | S | S | P |
|  | **E79 insertion** | - | E | E | E | E | E | E | E | E | E | E |
|  | R228S | R | S | S | S | S | S | S | S | S | S | S |
| *mtrR* | A39 | A | T | T | T | T | T | T | A | A | A | A |
|  | H105 | H | H | H | H | H | H | H | Y | Y | Y | H |
| *rplD* | G70D/S/A/R | G | D | Y | G | Y | G | G | G | G | G | G |
| Inferred from MIC | Ciprofloxacin | - | S | R | S | R | R | S | I | S | I | I |
| Inferred from MIC | Gentamycin | - | S | I | S | S | S | S | S | S | S | S |

We found six isolates carrying AMR mutations in penA C-terminal region (penicillin and cephalosporin resistance determinant) and four isolates with *ponA1* (penicillin and cephalosporin resistance determinant), respectively. We found the resistance determinants for penicillin-like D345 insertion and F504L mutation in *penA*^41^, we did not observe the cephalosporin resistance determinants. Five isolates harboured *gyrA* resistance mutations, two of which also had a *parC* mutation; while one strain had a mutation in only the *parC* gene. These mutations, in the Quinolone Resistance Determining Region (QRDR) of both the proteins, have been shown to contribute to the high MICs for ciprofloxacin^42^. Interestingly, one of the GyrA mutation we observed, D95A/G, and the mutation E91G in ParC, have been reported to be specific to Kenyan isolates^10^. Specific mutations in *porB1b* (penB AMR determinant), implicated in decreased influx of antimicrobials through the porin PorB^43,44^, were identified in five of the nine isolates. All nine strains harboured the V57M mutations in RpsJ, which has been shown to contribute to tetracycline resistance and R228S mutation in FolP which results in sulfonamide resistance^45^. Three strains harboured mutations in *rplD* gene, which is implicated in azithromycin resistance. Plasmid-mediated AMR determinants (highlevel resistance to benzylpenicillin and tetracycline -p*bla*TEM and p*tet*M) were detected in all strains except one where we detected only the TetM-containing plasmid. We did not identify *mef* or *erm* containing conjugative plasmids/transposons which are reported to cause resistance to macrolides^46^ or resistance-conferring mutations for spectinomycin.

### Understanding the basis of antibiotic resistance

We identified mutations in nine proteins associated with antibiotic resistance -GyrA, ParC, PonA1, RpsJ, PenA, PorB1b, FolP, RplD and MtrR. The mechanistic basis for resistance has been predicted for RpsJ (V57M) (tetracycline resistance), RplD (azithromycin resistance) and PenA mutations (Supplementary File). We investigated the basis of drug resistance for the protein mutations in GyrA, ParC, FolP, PonA1 and PorB1b (Supplementary File, Supplementary Figures 3a-c and 4a-c, template details in Supplementary Table 7) through homology modelling using templates with co-crystallized antibiotic structures. Models and alignments have been provided in Supplementary link.

*S. aureus* gyrase A and gyrase B dimer complexed with ciprofloxacin and DNA (PDB:2XCT)^47^ was taken as the template for modelling GyrA proteins from reference strain and strain 12 (F91 and G95). The substrates DNA and ciprofloxacin were retained in the modelled structure. The mutations S91F/Y and D95N/G/A have been reported to confer ciprofloxacin resistance. In the wild-type structure, the residue S91 formed polar contacts with the drug ciprofloxacin (Figure 2a,b). In the mutant, S91F, however, this contact was abolished (Figure 2c). We hypothesize that the mutation might result in the weakening of drug-protein interaction, thereby resulting in drug resistance.

**Figure 2:**
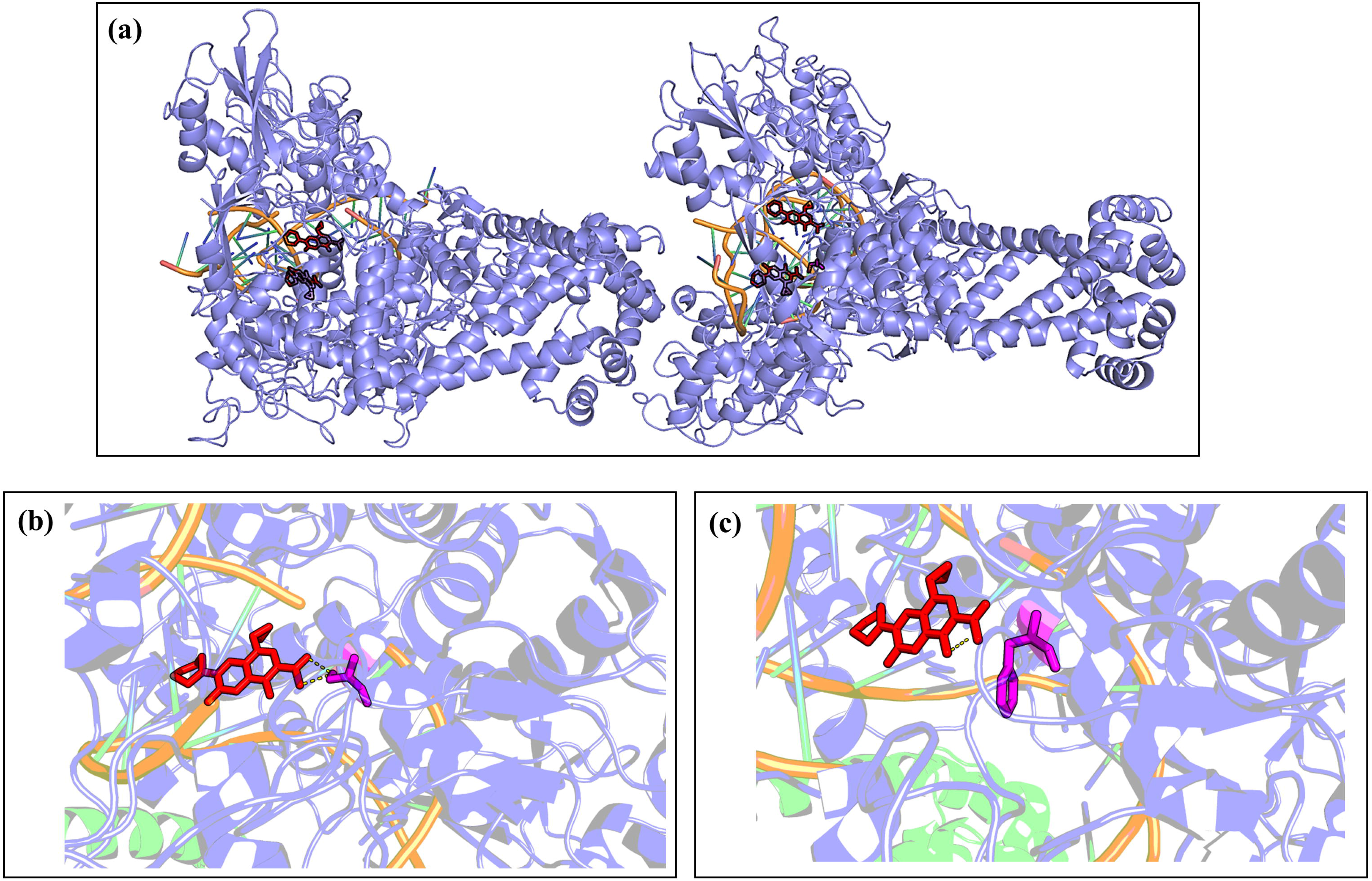
The structural basis of quinolone resistance mediated by GyrA. The modelled structure of the GyrA-GyrB complex with ciprofloxacin and DNA (a). A close-up of the antibiotic-binding region showing ciprofloxacin (red), S91 (magenta) in wild-type structure (b) and S91F mutant (magenta) (c). The polar contacts for the antibiotic are shown as yellow dashes.

ParC and ParE (DNA topoisomerase IV subunits A and B, respectively) are homologues of GyrA and GyrB^48, 49^. *A. baumannii* topoisomerase IV (ParE-ParC fusion truncate) cocrystallized with moxifloxacin and DNA (PDB id: 2XKK)^50^ was used as the template to model sequences from the reference strain and strain 12 (E91G). We observe that E91G mutation occurs close to S88, and these two residues are involved in a hydrogen bond network with the antibiotic, moxifloxacin (Figures 3a-c). E91 is also within 5Å of an Mg^2+^ ion which interacts with the antibiotic. In the case of the mutant, the hydrogen bonding network with the antibiotic is lost and the antibiotic interacts only with Mg^2+^. Thus, the interaction with moxifloxacin and other quinolones may be weakened in the case of the mutation.

FolP from *Yersinia pestis* crystallized with the substrate, 6-hydroxymethylpterin diphosphate and the sulfa drug, sulfamethoxazole (PDB id: 3TZF) was used as a template^45^ for modelling the wild-type and mutant FolP from the strain 3 (R228S mutation). Both R228 and S228 were seen to interact with the sulfa drug, the R228S mutation may interfere with the binding of the protein to the sulfa drug (Figures 4a-c).

**Figure 3:**
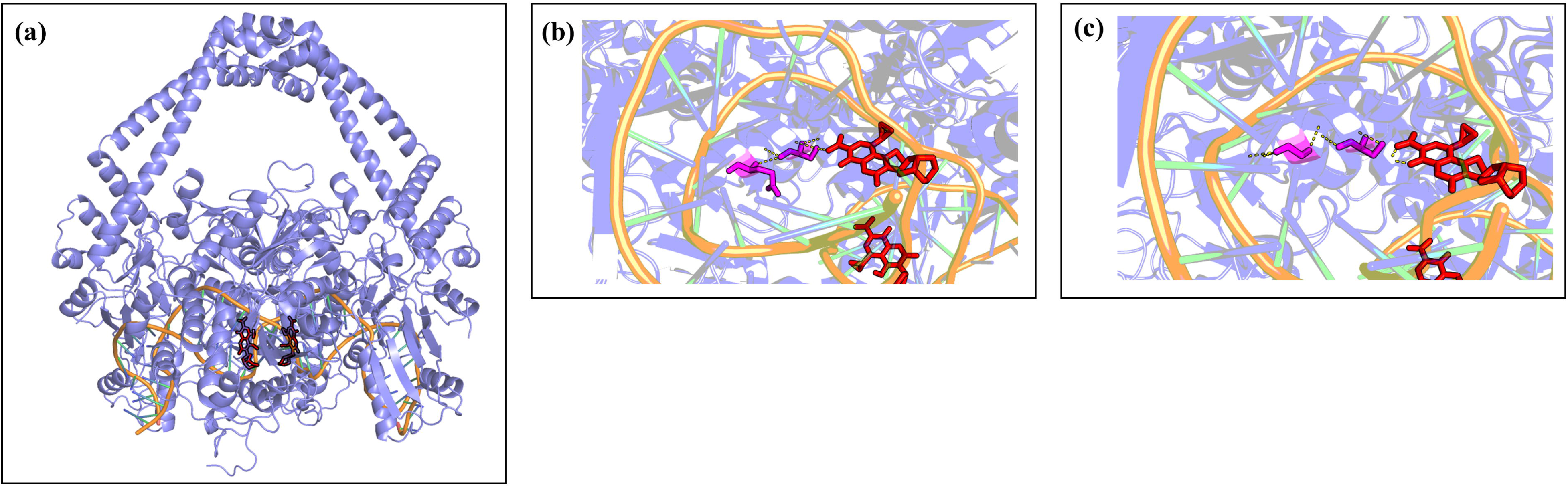
The mechanistic basis of quinolone resistance mediated by ParC. The modelled structure of the ParC-ParE complex with moxifloxacin and DNA (a). A close-up of the antibiotic-binding region showing moxifloxacin (red), S88 (magenta) and E91 in wild-type structure (b) and E91G mutant (magenta) (c). The polar contacts for the antibiotic are shown as yellow dashes.

### Phylogenetic relationship with other *N. gonorrhoeae* genomes from Kenya, other WHO geographical regions and WHO strains

The phylogenetic trees with Kenyan strains sequenced in this study and previously sequenced isolates (Cehovin et al and Kivata et al) revealed the presence of three clusters^10,17^. This is consistent with the three Sequence Type (ST) clusters derived by Cehovin et al using star-burst phylogeny. Our sequences clustered with Cluster 3, which has sequences from different STs (Figure 5a). Using a larger set of sequences from across the world, we obtained five clusters, with sequences from different geographical regions, co-clustering (Figure 5b). This is in contrast with a previous study reporting that Kenyan sequences clustered separately from sequences derived from UK and USA. The metadata for the strains like geographical region and antibiotic resistance details have been indicated in the phylogenetic trees (details in Supplementary Table 8).

**Figure 4:**
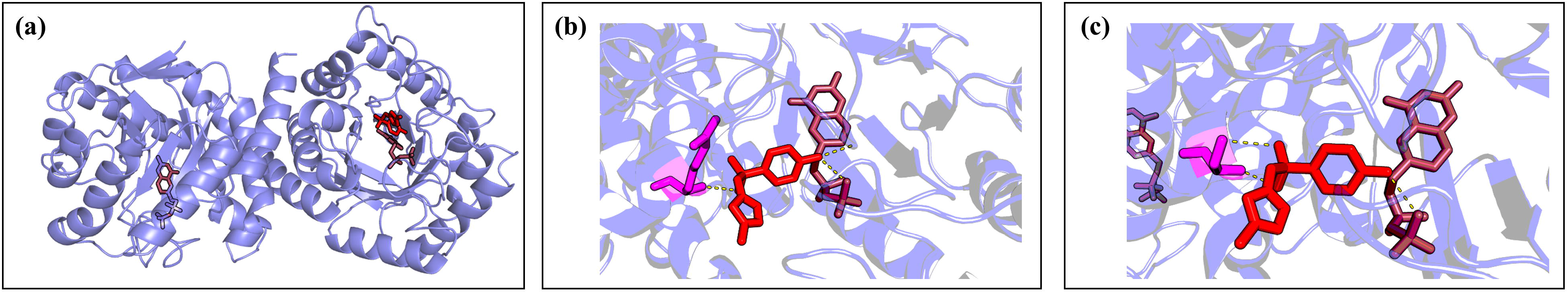
The mechanistic basis of quinolone resistance mediated by FolP. The modelled structure of the FolP complex with sulfamethoxazole and substrate, 6hydroxymethylpterin diphosphate (a). A close-up of the antibiotic-binding region showing sulfamethoxazole (red), the substrate (maroon) and R228 (magenta) in wild-type structure (b) and R228S mutant (magenta) (c). The polar contacts for the antibiotic are shown as yellow dashes.

**Figure 5:**
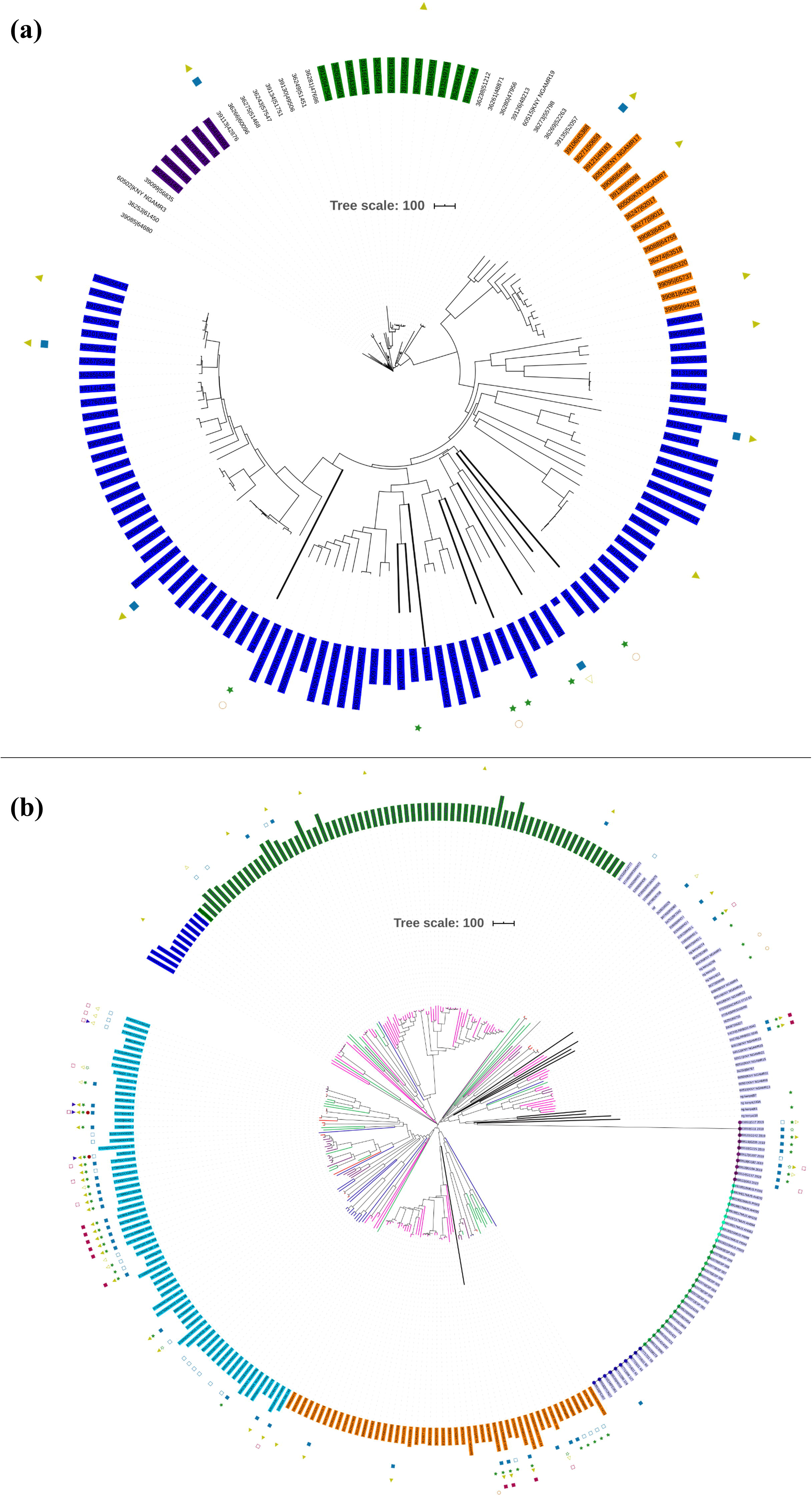
Phylogenetic tree of the sequenced strains with other Kenyan strains (a) and strains from other WHO geographical regions including WHO reference strains (b). The strains sequenced in this study have been indicated as black, bold nodes in the tree. The antibiotic resistance profile, wherever available, has been indicated as metadata (Penicillin-blue triangle, Ceftriaxone-maroon circle, Ciprofloxacin-dark green star, Tetracycline-light green triangle, Cefixime-dark blue triangle, Azithromycin-maroon circle, Gentamycin-orange circle. Filled shape-resistant, outline shape-intermediate resistant, no shape -susceptible). The labels for each clade as inferred from the phylogeny analysis has been indicated in different coloured labels. In (b) the strains from different geographical regions have been indicated in different colours (Africa-Green, Americas-Brown, Europe-Cyan, Western Pacific-Prussian blue, WHO-Red, Kenya-Magenta)

## Discussion

In the study, we present nine *N. gonorrhoeae* genomes sequenced using only the MinION sequencer to more than 98% coverage with high depth. The reference-based genome assembly obtained using bwa-mem was chosen for analysis, due to higher genome coverage and less number of indels and misassemblies compared to the reference genome. Although earlier studies have shown that bacterial MinION sequences show high mapping rates to the reference genome, with fewer indel rates, bwa-mem has found limited application with long error-prone reads^51–54^. We show that it is possible to obtain assemblies with good genome coverage for AMR determinant detection by using nanopore-only reads with a reference-based genome assembly, without depending on additional Illumina sequencing. Previous studies have obtained hybrid genome assembly for gonococcal isolates by combining short-read (Illumina) and long-read (ONT) sequence data^11,13^. This is another approach for genome assembly using MinION sequencing and, accordingly, an alternative to *de novo* assembly.

Databases like PubMLST, NGSTAR and PathogenWatch include profiles for resistance mediators like *penA*, *mtrR*, *penB*, *ponA*, 23S rDNA, *gyrA*, and *tetM* and do not include other determinants in their gonococcal AMR characterization module. We identified a set of AMR determinants from literature, including chromosomal gene mutations, promoter mutations, and the presence of plasmids with AMR genes. Plasmids containing genes encoding beta-lactamase and tetracycline resistance mediators (*tet*(M) and *bla*TEM) were identified in all but one strain (strain 12 contained only a TetM containing plasmid). This is consistent with a previous study that the plasmids are almost ubiquitous in Kenyan strains; thought to be because of the overuse of doxycycline for treating STIs^17^. Among the reported mutations, we observed S91F and D95Y in the QRDR (residues 55-110) of GyrA and E91G in the ParC QRDR (residues 66-119). We also observed the silent mutations in Y104 and L131 reported for ParC^12^. Novel mutations, V68A in the QRDR region of ParC (strain 285), E79 insertion in FolP and G70Y in RplD were also observed.

Based on homology modelling using templates crystalized with antibiotics, we see that the mutations in GyrA disrupt the hydrogen bonding with ciprofloxacin, and mutations in ParC occur very close to Ser 88 which is involved in hydrogen bondingwith moxifloxacin. We also observe that mutations in FolP at the substrate-binding site could affect the interaction with the antibiotic, as the residue at R228 interacts with sulfonamide. Phylogenetic study with previously sequenced Kenyan strains clustered our strains in the previously described cluster 3. From phylogenetic analysis using sequences from different countries, we observed five major clusters, with most of the Kenyan sequences (including the sequences from this study) cluster together with strains from different geographical regions.

## Conclusion

Using a literature mining approach, we show that it is possible to predict ciprofloxacin resistance using mutations in *gyrA*/*parC* in strains showing decreased susceptibility to ciprofloxacin. However, these AMR-associated mutations can also occur in susceptible strains, and molecular tests using gene-based PCR or WGS can be used to complement culture-based antibiotic resistance testing. Culture-based testing can reflect mutations in unknown antibiotic targets, but cannot predict if a patient can develop resistance later on, based on pre-existing mutations.

In conclusion, in the first reference-based genome assembly for gonococci using MinION, we show that using this approach we can obtain near-complete genomes that were effectively used for AMR and phylogenetic analysis. We also show that currently available tools for AMR analysis of gonococci are not able to capture many mutations listed in the literature. We have also provided a dataset of around 95 existing mutations in different genes implicated in AMR, plasmids, and efflux pumps which can be used by researchers across the world. This list will be continuously updated to keep up with the identification of new AMR targets/mutations. Here we demonstrate the potential of using MinION in resource-limited settings where NGS facility is unavailable. This can also be used in settings with concerns about the export of samples/DNA for WGS to other countries.

## Data Availability

The raw reads generated for this study have been deposited in BioProject Accession: PRJNA660404 (https://www.ncbi.nlm.nih.gov). The biosample details are available under the ids SAMN15960547, SAMN15960548, SAMN15960549, SAMN15960550, SAMN15960551, SAMN15960552, SAMN15960553, SAMN15960554, SAMN15960555. The genomes and annotation files are available under the bioproject.

## Author contributions

MJ collected the isolates, carried out the characterization, culturing and sequencing. AS, TD, AP and RK planned and carried out the sequencing. MI conceived and performed the bioinformatics analysis, and wrote the manuscript with inputs from SK, MJ, MM and AS. RN, PM, MM, RC, JN and JG performed the experiments. SK, RS and OA provided intellectual support and valuable discussions. All authors listed have made a substantial contribution to the work and approved it for publication.

## Conflicts of interest

The authors declare that they have no competing financial interests that could have influenced the work reported in this paper.

## Ethics approval

The ethics approval for the study was obtained from the Kenyatta National Hospital University of Nairobi Ethics and Research Committee, Nairobi, Kenya, for the use of anonymized samples collected from female patients attending local Sexually Transmitted Disease (STD) clinics.

## Acknowledgements

This work was supported by the Indo Africa capacity component of the dengue sequencing to vaccine grant from Mr. Narayana Murthy (Infosys) and the NCBS core funds to SK. The authors would like to acknowledge KAVI-ICR for the initial funding for the project. We thank NCBS (TIFR) and KAVI-ICR for infrastructural and financial support.

## Notes

### Competing Interest Statement

The authors have declared no competing interest.

### Author Declarations

Kenyatta National Hospital - University of Nairobi Ethics and Research Committee, Nairobi, Kenya

